# Altered Functional Networks during Gain Anticipation in Fibromyalgia

**DOI:** 10.1101/2023.04.28.23289290

**Authors:** Su Hyoun Park, Andrew M. Michael, Anne K. Baker, Carina Lei, Katherine T. Martucci

**Affiliations:** Department of Anesthesiology, Human Affect and Pain Neuroscience Laboratory, Duke University School of Medicine, Durham, NC, USA; Center for Translational Pain Medicine, Duke University Medical Center, Durham, NC, USA; Duke Institute for Brain Sciences, Duke University, Durham, NC, USA

**Keywords:** chronic pain, fibromyalgia, gain anticipation, motor network, monetary incentive delay (MID) task

## Abstract

Reward motivation is essential in shaping human behavior and cognition. Previous studies have shown altered reward motivation and reward brain circuitry in chronic pain conditions, including fibromyalgia. Fibromyalgia is a chronic disorder characterized by widespread musculoskeletal pain, fatigue, cognitive problems, and mood-related symptoms. In this study, we analyzed brain reward networks in patients with fibromyalgia by using a data-driven approach with task-based fMRI data. fMRI data from 24 patients with fibromyalgia and 24 healthy controls were acquired while subjects performed a monetary incentive delay (MID) reward task. Functional networks were derived using independent component analysis (ICA) focused on the gain anticipation phase of the reward task. Functional activity in the motor, value-driven attention, and basal ganglia networks was evaluated during gain anticipation in both patient and healthy control groups. Compared to controls, the motor network was more engaged during gain anticipation in patients with fibromyalgia. Our findings suggest that reward motivation may lead to hyperactivity in the motor network, possibly related to altered motor processing, such as restricted movement or dysregulated motor planning in fibromyalgia. As an exploratory analysis, we compared levels of motor network engagement during early and late timepoints of the gain anticipation phase. Both groups showed greater motor network engagement during the late timepoint (i.e., closer to response), which reflected motor preparation prior to target response. Importantly, compared to controls and consistent with the initial findings described above, patients exhibited greater engagement of the motor network during both early and late timepoints. In summary, by using a novel data-driven ICA approach to analyze task-based fMRI data, we identified elevated motor network engagement during gain anticipation in fibromyalgia.

## INTRODUCTION

Reward motivation is an essential factor that shapes human behavior and cognition (Banich & Floresco, 2019). The accurate processing of rewarding events directly affects an individual’s maximized gains and minimized losses (Kahneman & Tversky, 2013) such that reward processing is crucial for survival. Reward processing involves learned associations between reward and stimuli (e.g., monetary reward) which guide human cognitive processes, such as attention, memory, and decision-making (Daw & Doya, 2006). Accordingly, during reward anticipation cognitive processes exhibit different patterns (Chiew et al., 2016). For example, during reward anticipation, cognitive-motor and cognitive-attention networks activate to prepare for achieving a reward (Critchley et al., 2001; Ivanov et al., 2012; Pornpattananangkul & Nusslock, 2015).

Given their essential role in shaping human cognition and behavior, brain reward systems have been extensively investigated using neuroimaging of clinical populations. Specifically, brain reward systems are critical in modulating pain experiences (Becerra et al., 2001; Becerra & Borsook, 2008; DosSantos et al., 2017) and consistently appear to be dysregulated in patients with chronic pain conditions such as fibromyalgia (Berger et al., 2014; Martucci et al., 2018, 2019; Park, Baker, et al., 2022). Fibromyalgia is a chronic pain condition characterized by widespread musculoskeletal pain that is frequently accompanied by fatigue, cognitive problems, and mood-related symptoms (Sluka & Clauw, 2016). When performing the monetary incentive delay (MID) reward task, patients with fibromyalgia exhibit altered medial prefrontal cortex (MPFC) activity during gain anticipation (Martucci et al., 2018). In another cohort of patients with chronic low back pain and fibromyalgia, Kim et al. (2020) used a MID task and revealed that patients had a decreased response in the right striatum to reward outcomes.

As revealed by functional magnetic resonance imaging (fMRI) studies using a measure of functional connectivity (Biswal et al., 1995), corticostriatal circuits (i.e., MPFC – nucleus accumbens functional connectivity) are dysregulated in patients with chronic back pain (Baliki et al., 2010, 2012) and fibromyalgia (Park, Baker, et al., 2022). Such alterations in corticostriatal reward circuits may relate to dysregulated reward processing which, in turn, may be a crucial contributor to chronic pain (Navratilova & Porreca, 2014). Thus, by examining how reward processing is represented in brain networks in patients, we may improve the understanding of how neural mechanisms contribute to chronic pain.

While most research investigating brain reward circuits in chronic pain have used functional connectivity analyses of resting-state fMRI data (Baliki et al., 2010; Park et al., 2022), very few studies have used functional connectivity analyses of task-fMRI data acquired while participants performed a reward task inside the MR scanner (Park, Deng, et al., 2022). Meanwhile, as consistently demonstrated by several reward based task-fMRI studies, patients with chronic pain show altered brain reward response to gain anticipation (Baker et al., 2022; Kim et al., 2020; Loggia et al., 2014; Martucci et al., 2018; Park, Deng, et al., 2022). However, in contrast to such analyses which focus on single brain regions or isolated functional connections, to our knowledge neuroimaging studies have not investigated functional network-level alterations during gain anticipation among patients with chronic pain.

In this study, we investigated brain reward processes in patients with chronic pain vs. healthy individuals during gain anticipation, with a novel focus on activity within pre-defined functional networks. As described above, many prior neuroimaging studies provide evidence of altered brain reward circuits in fibromyalgia (Loggia et al., 2014; Martucci et al., 2018; Park, Baker, et al., 2022). To compute overall measures of reward-associated function within defined brain networks, we used a data-driven approach—specifically, independent component analysis (ICA). Importantly, ICA does not require selection of *a priori* brain regions and makes no assumptions about the hemodynamic response function and helps to uncover novel altered connectivity patterns using a data-driven approach (Bhaganagarapu et al., 2013; Calhoun et al., 2001; McKeown et al., 1998; Ray et al., 2013).

Before performing ICA, we identified three functional networks that are related to gain anticipation: 1) the basal ganglia network, 2) the value-driven attention network, and 3) the motor network. As demonstrated by numerous previous studies, brain subcortical regions (e.g., nucleus accumbens (NAcc), caudate, and putamen) play a crucial role in processing reward stimuli (Knutson, Adams, et al., 2001; Knutson et al., 2000; Russo & Nestler, 2013; Thut et al., 1997). Additionally, as revealed by clinical neuroimaging studies, basal ganglia responses to gain anticipation are dysregulated in individuals with severe institutional deprivation early in life (Mehta et al., 2010) and childhood adversity (Dillon et al., 2009). The value-driven attention network was first introduced by Anderson (2017) and the primary brain regions included in this network are the visual-corticostriatal loop, including the lateral occipital cortex (LOC), early visual cortex, caudate tail, and intraparietal sulcus (IPS). Increased activity in the value-driven attention network has been shown in response to high-value reward (Anderson, 2017). In addition, increased activity within components of attention networks, specifically in the inferior parietal and lateral occipital cortices, are observed during gain anticipation in hybrid tasks based on the MID task (Ivanov et al., 2012). Finally, as observed in reward-related task-based fMRI studies that used the MID task, motor cortex activity is enhanced during gain anticipation specifically when preparing motor response to earn reward (Knutson, Fong, et al., 2001). Thus, we focused our analysis on the motor, attention, and basal ganglia networks. In addition, we visually inspected the salience network and the default mode network (DMN) to confirm that ICA accurately extracted the functional networks and to perform additional supplementary analysis. For each of these networks, we tested the association between the timecourse of gain anticipation of our task presentation and the timecourse of the ICA-derived brain network. We then compared the degree of task-to-network correlation between patients with fibromyalgia versus healthy controls to identify group functional differences. Lastly, in an exploratory analysis, we compared the brain network activity during early vs. late gain anticipation.

## PARTICIPANTS AND METHODS

### Participants

Twenty-six patients with fibromyalgia and 28 healthy controls participated in the study. All study procedures were approved by the Duke University Institutional Review Board. Patients with fibromyalgia were required to meet modified American College of Rheumatology 2016 criteria for fibromyalgia (Wolfe et al., 2016). Patients reported an average pain score of at least 2 (0–10 verbal scale) in the previous month and had pain in all four quadrants of the body. No patients reported uncontrolled depression or anxiety.

### Medication Usage

Patients were allowed and expected to continue regular use of their medications during the study. All patients were not taking opioid medications at the time of the study, reported no opioid use for more than 90 days prior to the study, and reported no prior opioid use for longer than 1 month in their lifetime. Out of a total 26 patients included in this study, 25 patients were taking one or more of the following medications: nonsteroidal anti-inflammatory drugs (NSAIDs; N = 7), acetaminophen (N = 5), other pain medicine (e.g., linaclotide, salicylate; N = 2), serotonin-norepinephrine reuptake inhibitors (SNRIs; N = 7), selective serotonin reuptake inhibitors (SSRIs; N = 5), other anxiolytics (e.g., lurasidone; N = 1), triptans (N = 5), trazodone (N = 2), antiepileptic (N = 2), muscle relaxants (N = 7), gamma-aminobutyric acid (GABA) analogs (e.g., pregabalin and gabapentin; N = 6), benzodiazepine (N = 7), norepinephrine–dopamine reuptake inhibitor (NDRI; N = 4). One patient reported not taking any pain or mood-altering medications. All 28 healthy control participants reported not taking any pain or mood-altering medications. However, 2 of the controls reported taking single doses of medications for menstrual pain and a sleep aid in the weeks prior to the study visit (NSAIDs, N = 1; benzodiazepine, N = 1). We tested whether the exclusion of these two healthy control datasets would influence our group results, and as exclusion did not significantly change group results, these controls were included in the final analysis.

### Study Procedures

All study procedures were conducted at the Duke-UNC Brain Imaging and Analysis Center. Before scanning, participants received instructions on the monetary incentive delay (MID) task and performed a short practice session. Functional scans were acquired during two runs of the MID task (each approximately 11-12 minutes duration). During each MID task run, 6 variations of trials were presented with the following monetary cues: +$5.00, +$1.00, +$0.00 (high-, low-, no-gain); –$5.00, –$1.00, –$0.00 (high-, low-, no-loss). The first run of the MID task consisted of 42 trials. The second run of the task consisted of 48 trials. Each trial consisted of 4 consecutive presentations: a cue, fixation cross, triangle target, and feedback. Additional information regarding trial timing is described in Figure 1. To achieve a participant’s hit rate of approximately 66% across trials (Knutson et al., 2005), the target duration was automatically adjusted based on each participant’s performance. The MID task was programmed in MATLAB with Psychophysics Toolbox v.3 (Brainard, 1997; Kleiner et al., 2007). In the current study, as described in the introduction, our primary research question was focused on the gain anticipation phase (i.e., for +$5 and +$1 trials only; see highlighted yellow background in Figure 1).

**Figure 1.**
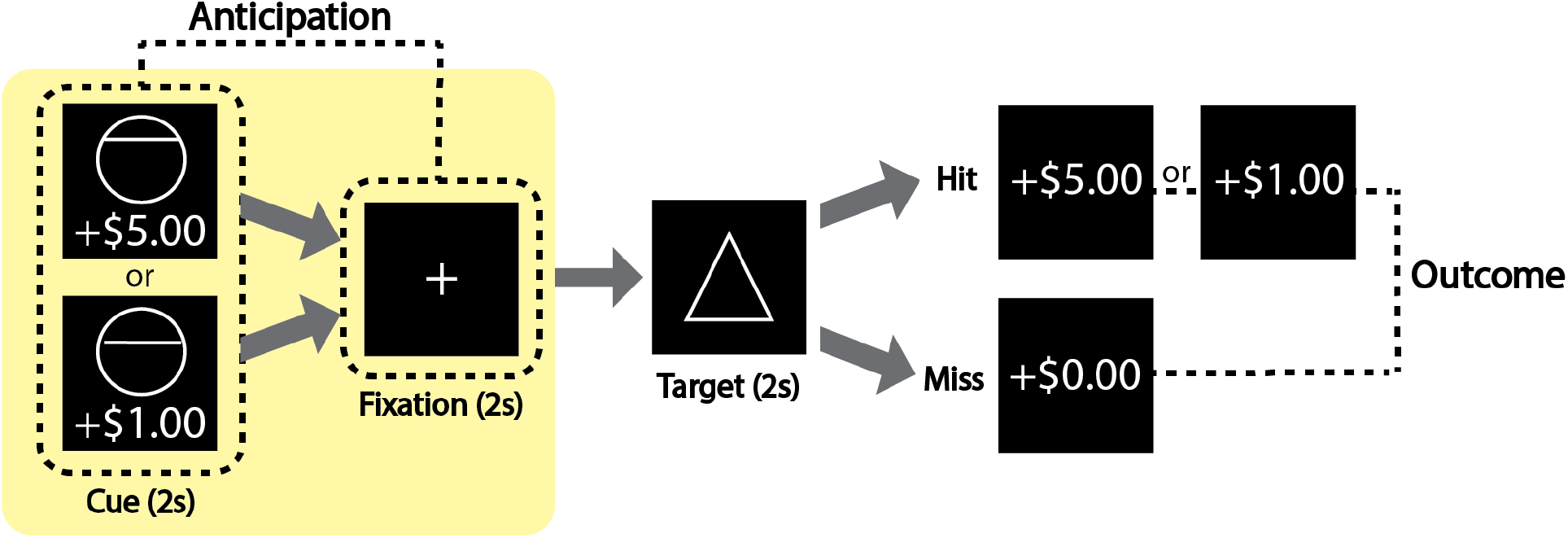
Gain trials of the monetary incentive delay (MID) task. Each trial consisted of an anticipation phase and an outcome phase of gain trials (loss and neutral trials were not analyzed in this study). Each trial (TR-locked; TR=2 seconds) consisted of 4 TRs with each TR corresponding to the cue, fixation, target, and outcome. After each trial was a variable-duration inter-trial interval for 5∼7 TRs. Cues were either circles (potential gain trials) or squares (potential loss trials; not described in this figure). Cues were presented with monetary values (for gain trials: +$1 and +$5). Note: Other monetary values were presented during the task but were not used in this study (for loss trials: -$1 and -$5; neutral trials $0). After a fixation period, a target period began. A triangle was presented for a variable duration (∼250 ms) during the target period; the duration of triangle “target” presentation was determined on prior response accuracy and adjusted throughout the task to obtain an average 66% hit rate. During the outcome phase, hit or miss (i.e., win or no win) feedback was given. After the feedback, a black screen was presented with a pseudo-randomized inter-trial interval period of 1-3 TRs (2, 4, or 6 s duration).

### MRI Data Acquisition

All scans were acquired on a 3T GE Premier UHP system with a 48-channel head coil at the Duke-UNC Brain Imaging and Analysis Center. The scans consisted of the preparatory localizer, calibration, two MID task functional scans, and a T1 anatomical scan. Functional images for multi-band (factor = 3) scan parameters were as follows: flip angle 76°, echo time (TE) 30 ms, repetition time (TR) 2 seconds, 69 slices, 2 mm slice thickness, pixel size 2 mm. The two MID task scans consisted of 266 and 302 volumes, respectively, and we excluded the first 4 lead-in volumes (8 seconds) for scanner stabilization. The T1 MPRAGE anatomical scan parameters were as follows: field of view (FOV) 256 mm, frequency direction anterior/posterior, flip angle 8°, TE 3.2 ms, TR 2.2 seconds, 1 mm slice thickness.

### MRI Data Preprocessing

The CONN Toolbox v.20b (Whitfield-Gabrieli & Nieto-Castanon, 2012) running on MATLAB v.R2019a and SPM12 were used for preprocessing functional data. The default preprocessing pipeline was used. Functional data underwent realignment, and slice-timing correction was manually applied using the multi-band timing information. Outliers were identified using the CONN toolbox’s intermediate outlier identification setting (i.e., framewise displacement above 0.9 mm or global blood oxygenation level-dependent signal changes above 5 standard deviations were identified as outliers and automatically excluded from the analysis). Structural and functional images were segmented into three tissue classes (grey matter, white matter, and cerebrospinal fluid) and normalized to Montreal Neurologic Institute (MNI) space. Spatial smoothing with 4 mm full width half maximum was applied. Then, CONN’s default denoising pipeline was used. Band-pass filtering to 0.008-0.09 Hz was applied, and estimated subject-motion parameters were generated to further regress out motion parameters from each participant’s functional network timecourse.

### Group Independent Component Analysis

A group independent component analysis (GICA) was performed on the functional data for the two MID task scans using GIFT v4.0 software running in MATLAB R2019a (Calhoun et al., 2001). GICA computes brain functional networks and their timecourses in a data-driven manner. The denoised preprocessed functional data (from the two MID scan sessions from all subjects) were provided as input into the GICA toolbox. The fMRI data were decomposed into 30 functional networks using the InfoMax algorithm and were visually inspected. Brain regions within the ICA networks were manually labeled using previously established functional networks from the Harvard Oxford MNI atlas (Desikan et al., 2006). Networks of interest were selected based on their relevance to reward processing and included: the motor network (with a focus on the left motor network as all participants were instructed to respond to the task using their right index finger), basal-ganglia network, and value-driven attention network for the main analysis, and salience network and DMN for the exploratory analysis.

### Functional Network Activation During Gain Anticipation

Patients with fibromyalgia vs. healthy controls group differences were evaluated for each functional network’s temporal correlation with gain anticipation during task presentation (Figure 2). First, we modeled the timecourse of gain anticipation using task presentation timing. Using the FMRIB Software Library (FSL; Jenkinson et al., 2012), we constructed the task model timecourse by convolving the timecourse of the anticipation phase with the hemodynamic response function. Then, we correlated the task timecourse with the timecourse of each functional network produced by ICA. At this step, to rigorously reduce the impact of motion, we used MATLAB’s multilinear regression function (“regstats”) to regress out six motion parameters (three translation and three rotation) from each participant’s functional network timecourse. We repeated this process for all participants. Finally, we performed patients with fibromyalgia vs. healthy controls group comparison (for detailed description, see Supplementary Figure 1).

**Figure 2.**
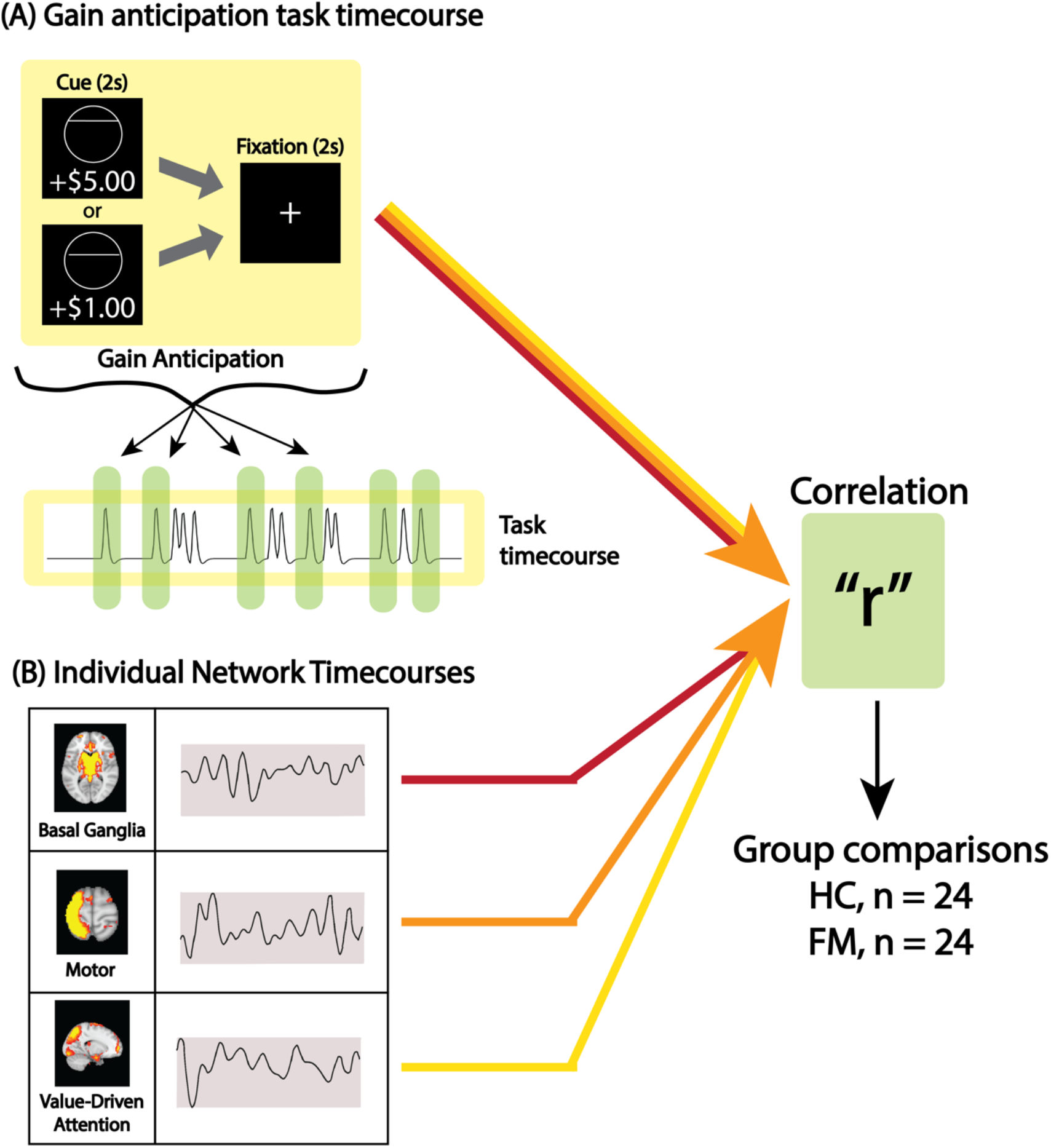
Example pipelines for the analyses of functional network activation during gain anticipation. (A) Gain anticipation task timecourses constructed by convolving the timecourse of the MID task anticipation phases with the hemodynamic response function. (B) Functional ICA network timecourses resulting from the ICA analysis. Correlations were evaluated between the task timecourse (A) and each of the 3 network timecourses for the main analysis (B) at the individual-level. Then, individual correlation values were averaged for each group to evaluate group differences. Abbreviation: FM, fibromyalgia; HC, healthy control; r, Pearson correlation coefficient.

### Additional Analyses

As an exploratory analysis, we measured changes in the degree of activation in each functional network at different timepoints during the gain anticipation phase. For this analysis, we divided the gain anticipation phase into two distinct timepoints: “early” and “late” and repeated the same analysis steps as described above. For this exploratory analysis, the task timecourses were constructed by convolving with the hemodynamic response function the timecourses of the 1) early timepoint (during cue presentation) and 2) late timepoint (during fixation presentation).

### Statistical Analysis

Group differences in reaction time and accuracy rate were evaluated using an unpaired two-sample t-test. Group differences for the selected functional networks were evaluated for the gain anticipation task timecourse. After performing correlation analysis between each functional network timecourse and the gain anticipation task timecourse, we used the resulting Pearson’s R values to evaluate group differences in functional network activation using a two-sample t-test. For the main analysis, Bonferroni correction was applied to identify a corrected P-value threshold (i.e., correction for three brain networks; p = 0.016). For the exploratory analyses, we followed the same statistical analysis steps as described above for the main analysis, but results were not corrected for multiple comparisons.

## RESULTS

### Participant Demographics

Twenty-six patients with fibromyalgia and 28 healthy controls participated in the study. Two patients and four controls were excluded due to excessive head motion (max motion above 3mm; identified as outliers in CONN as their max motion was above the third quartile +1.5 × interquartile range [IQR]). There was no significant age difference between patients and healthy controls (patients: M = 37.87, SD = 13.89; controls: M = 41.79, SD = 12.36). The duration of fibromyalgia-related pain symptoms reported by patients ranged from 9 months to 20 years (M = 8.58, SD = 8.50). Demographic data is provided in **Table 1**.

**Table 1.** Participant Demographics. No participants were of race categories for American Indian, Pacific Islander, Alaskan, or Native American. “Other” refers to self-identified race other than the pre-defined categories.

|  | Patients | Controls |
| --- | --- | --- |
| **Total Participants**<br>**(all female)** | **24** | **24** |
| **Righthanded** | **23** | **22** |
| **Self-Identified Race<sup>a</sup>** |  |  |
| **Asian** | **0** | **3** |
| **Caucasian** | **18** | **19** |
| **African American** | **5** | **2** |
| **Other** | **1** | **0** |
| **Hispanic or Latina Ethnicity** | **1** | **0** |
| **Employment Status** |  |  |
| **Part-time employed** | **3** | **2** |
| **Full-time employed** | **13** | **16** |
| **Unemployed** | **8** | **6** |
| **Income Level<sup>b</sup>** |  |  |
| **\$0–\$34,999** | **11** | **4** |
| **\$35,000–\$59,999** | **7** | **7** |
| **\$60,000 or more** | **6** | **11** |
| **Education Level** |  |  |
| **High School** | **1** | **0** |
| **College/University** | **21** | **16** |
| **Advanced Degree** | **2** | **8** |

<sup>a</sup> One patient self-identified with two racial categories.
<sup>b</sup> Two controls did not report their income level.

### Behavioral Measures: Reaction Time and Accuracy Rate

Reaction time was not collected from one patient; as such, data from 23 patients and from 24 controls were included in the reaction time analysis (i.e., one patient did not respond to the loss trials with -$1, -$5, and +$0 cues). However, we did not exclude this patient’s data from the main analysis which only focused on gain trials. Reaction time was faster for gain and loss trials compared to no gain/no loss trials [F(5,225) = 3.9, p = 0.002]. However, for reaction time, we identified no group effect [F(1,225) = 0.2, p = 0.694] and no interaction (group x trial conditions) [F(5,225) = 0.3, p = 0.928].

As in prior studies (Knutson et al., 2008; Martucci et al., 2018), the MID task was programmed to track participant’s performance by automatically changing the required response time to target 66% accuracy rates on average. Percent hits were greater for gain and loss trials compared to no gain/no loss trials [F(5,230) = 7.6, p < 0.001]. However, no group effect [F(1,230) = 0.4, p = 0.542] or interaction effects (group x trial conditions) [F(5,230) = 0.5, p = 0.745] were observed. There were no significant group differences in the behavioral measures, suggesting that patients and controls were similarly engaged in the task.

### Functional Networks

Functional network analyses focused on the following three networks of interest: basal-ganglia, value-driven attention, and left motor networks. These networks are activated during reward processing (Anderson, 2017; Knutson et al., 2000; Knutson, Fong, et al., 2001). We visually inspected ICA results based on spatial overlap with standard network examples (Anderson, 2017; Biswal et al., 1995; Di Martino et al., 2008) and identified that, among the 30 total functional networks, three closely resembled our networks of interest. As shown in Figure 3, the left motor network includes the left superior and middle frontal gyrus and the left pre- and post-central gyrus. The value-driven attention network includes the early visual cortex, lateral occipital cortex, intraparietal sulcus, and caudate tail. The basal-ganglia network includes a large portion of subcortical brain regions such as the NAcc, thalamus, caudate, pallidum, hippocampus, amygdala, and putamen.

**Figure 3.**
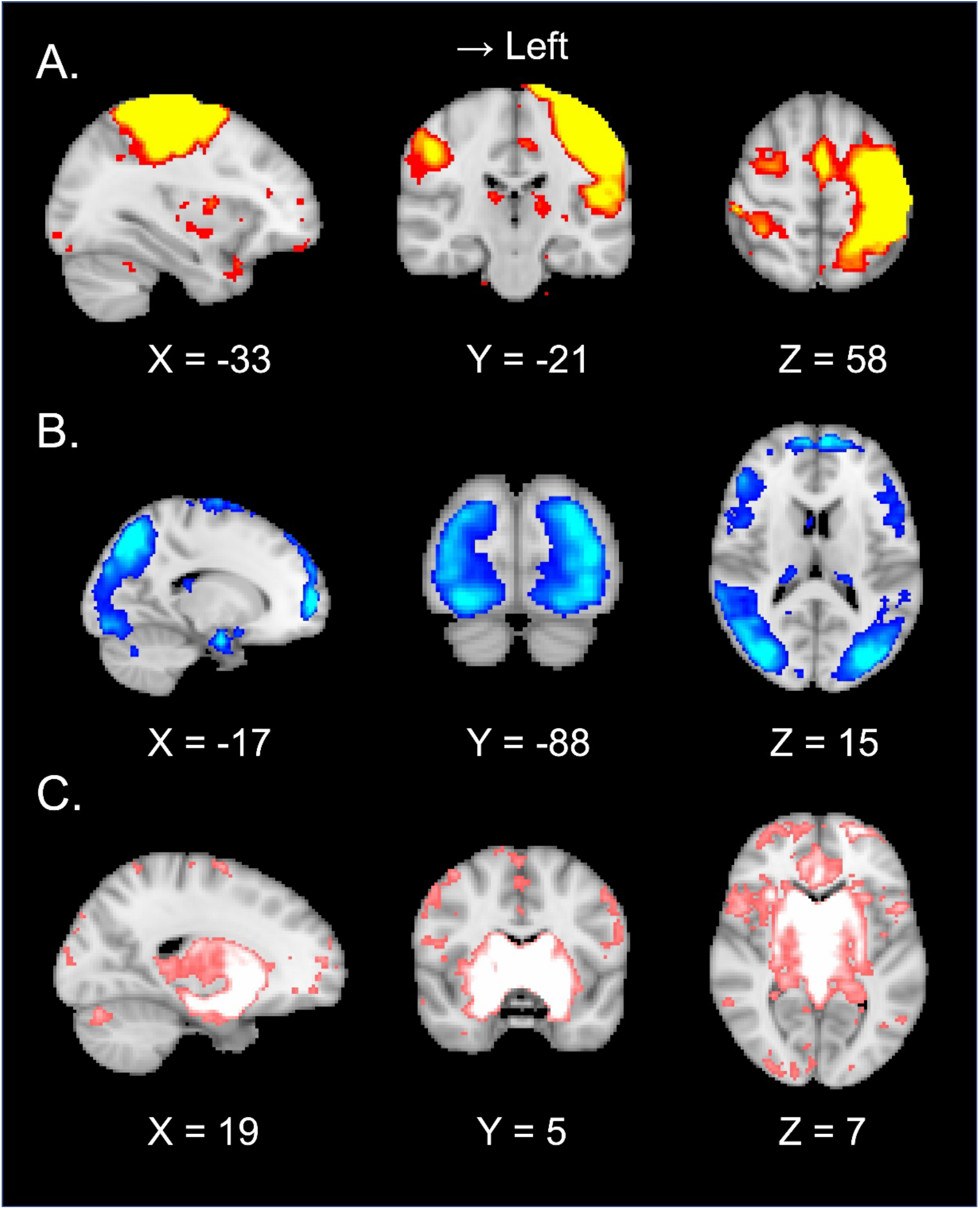
Functional networks extracted during MID task performance. Functional networks are shown at peak activation with MNI coordinates. (A) The left motor network includes the left superior and middle frontal gyrus and the left pre- and post-central gyrus. (B) The value-driven attention network includes the early visual cortex, lateral occipital cortex, intraparietal sulcus, and caudate tail. (C) The basal-ganglia network includes a large portion of subcortical brain regions such as the NAcc, thalamus, caudate, pallidum, hippocampus, amygdala, and putamen. Abbreviation: MID, monetary incentive delay; MNI, Montreal Neurological Institute; NAcc, nucleus accumbens.

From the total 30 functional networks identified by ICA, additional networks that resembled pre-established networks (e.g., default mode network, salience network) are presented in Supplementary Figure 2.

### Group Differences in Correlation of Functional Network Activation to Gain Anticipation

Each of our 3 functional network timecourses were positively, but not significantly, correlated with the gain anticipation task timecourse [left motor network: r = 0.10, p = 0.21; value-driven attention network: r = 0.07, p = 0.28; basal-ganglia network: r = 0.10, p =0.22]. This result was expected because in contrast to the gain anticipation timecourse, the functional network timecourses (i.e., ICA-derived data) contained data from all task phases (e.g., gain anticipation, loss anticipation, reward outcome phase). Nevertheless, as described in the introduction, our research question focused on whether functional networks would be altered in patients with fibromyalgia during gain anticipation.

As revealed by the group network analysis, the left motor network had a significantly stronger correlation in patients vs. controls [t(46) = 2.75, p = 0.008; Fig. 4A]. In contrast, the value-driven attention network and basal-ganglia network were not significantly different between groups (p = 0.13 and p = 0.55, respectively; Fig. 4B and 4C). The results of our exploratory analyses (i.e., the DMN and salience network) are described in Supplementary Figure 2.

**Figure 4.**
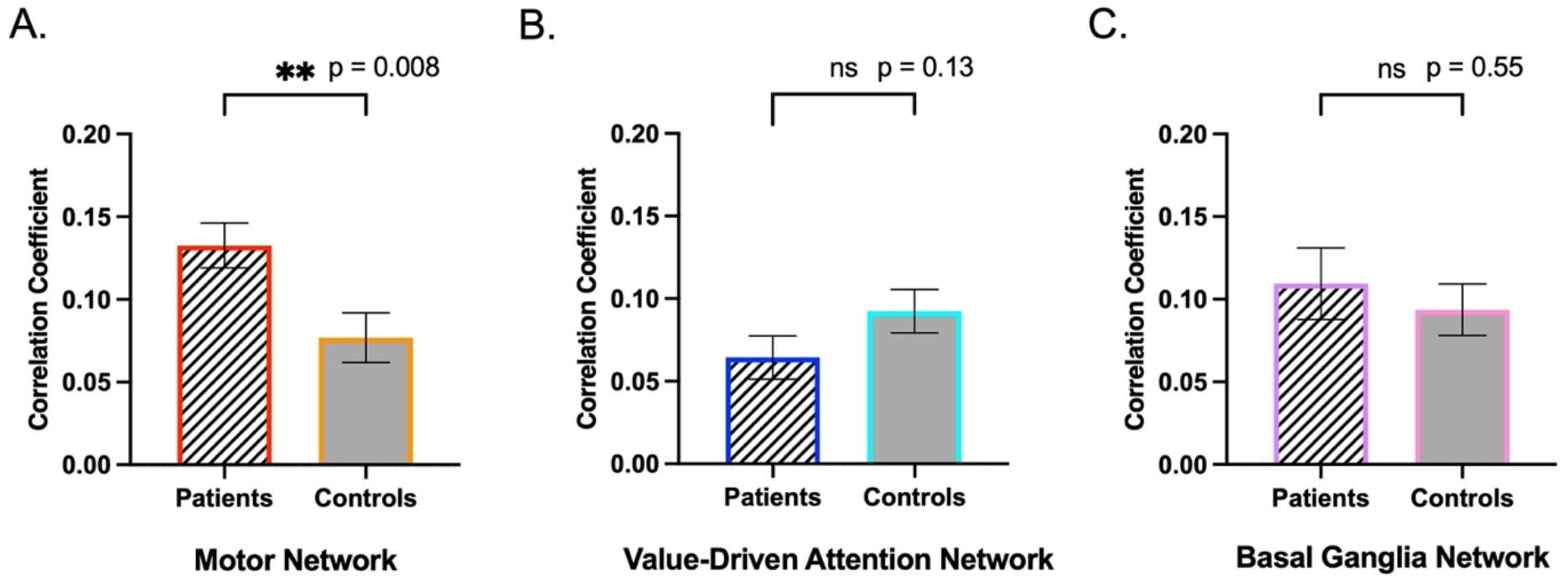
Correlation between gain anticipation task presentation and activation in (A) left motor network, (B) value-driven attention network and (C) basal ganglia network. Bar line colors are matched with the brain image color in Figure 3.

### Exploratory Analysis: Motor Network Activity During Early and Late Timepoints of the Gain Anticipation Phase

In the primary analysis, the task timecourse for the gain anticipation phase was modeled with 4 seconds (including both cue and fixation presentations). Based on identified significant group differences in the motor network’s relationship to gain anticipation, we divided the gain anticipation phase into two timepoints: “early” (cue presentation; 2 seconds) and “late” (fixation presentation; 2 seconds) (see “Anticipation” phase in Figure 1). Then, separately for early and late timepoints, we modeled each task timecourse and correlated it with the timecourse of functional network. By performing this separate analysis, we examined the potential effect of temporal sequence in representation of functional networks during gain anticipation.

Using two-way analysis of variance (ANOVA) [presentation (early vs. late) x group (patients vs. controls)], we measured correlation values across different presentation times and groups. For the motor network, we identified a significant main effect of presentation time [F(1, 92) = 8.22, p = 0.005], suggesting that correlations increased over time. We also found significant group effects [F(1, 92) = 13.11, p < .001), such that patients showed significantly greater correlations across both presentation times [post-hoc t-test; early timepoint: t(46) = 2.77, p = 0.007; late timepoint: t(46) = 2.33, p = 0.024]. No interaction effect was observed (Fig. 5A). For the value-driven attention network, we identified a significant main effect of presentation time, F(1, 92) = 12.65, p < 0.001, and a trend of main effect of group, F(1. 92) = 3.85, p = 0.053. As revealed by post-hoc t-test, a trend group difference was identified only in the early timepoint, t(46) = -1.79, p = 0.079. No interaction effect was observed (Fig. 5B). For the basal ganglia network, we identified a significant main effect of presentation time, F(1, 92) = 11.70, p < 0.001, but no group and no interaction effects were observed (Fig. 5C).

**Figure 5.**
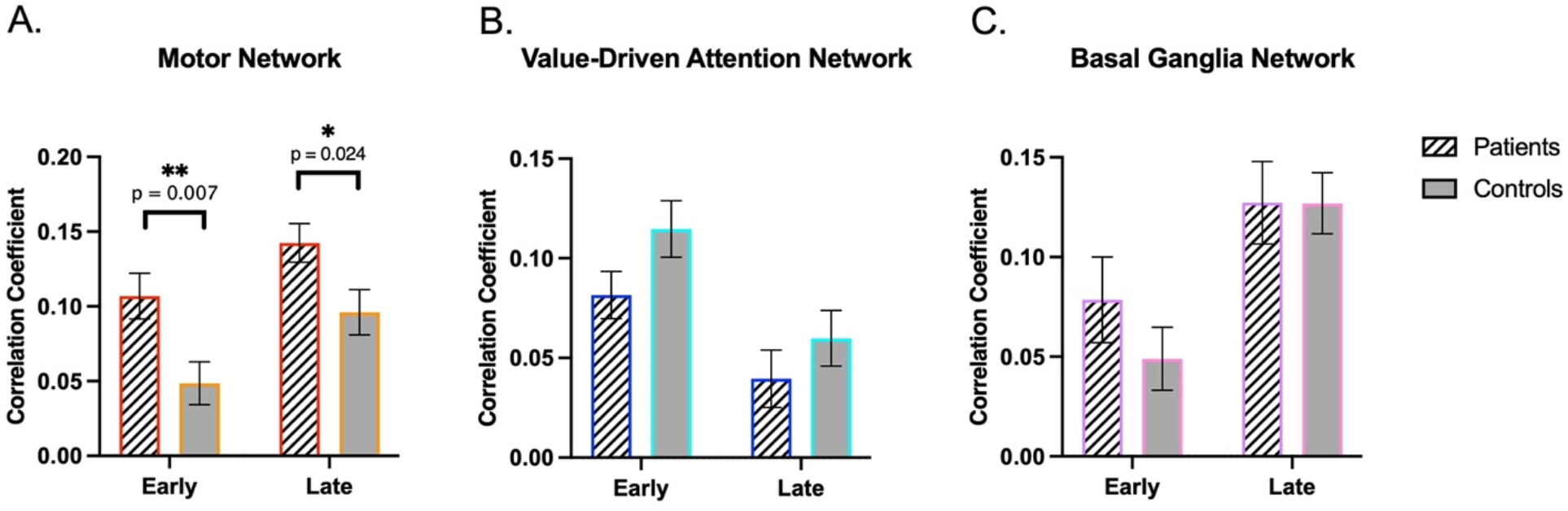
Group differences in functional network – gain anticipation correlation coefficients during early vs. late timepoints. (A) For the left motor network, patients showed higher correlations with the task timecourse than controls at both early and late timepoints of gain anticipation. In addition, the correlations were higher during the late timepoint in both groups. (B) As only trend-level effects, for the value-driven attention network, correlations with the task timecourse were stronger (not significant) during the early timepoint of gain anticipation than during the late timepoint, and the correlation was stronger (not significant) in the control group than in the patient group. (C) For the basal ganglia network, the correlation with the task timecourse was stronger during the late timepoint than during the early timepoint in both groups. Bar line colors are matched with the brain image color in Figure 3.

## DISCUSSION

For the first time, by using a data-driven ICA approach to analyze reward task fMRI data, we aimed to determine how functional networks are engaged during gain anticipation, and how this is altered in patients with fibromyalgia. Specifically, we used a new set of multi-band fMRI data that was collected from patients with fibromyalgia and healthy controls while they performed a MID task. For our networks of interest, we focused on the basal-ganglia, value-driven attention, and left motor networks, as they activate during the anticipation of reward (Anderson, 2017; Knutson et al., 2000; Knutson, Fong, et al., 2001). As compared to controls, patients with fibromyalgia demonstrated significantly higher engagement in the motor network during gain anticipation. In a follow-up exploratory analysis, we divided the anticipation phase into two timepoints — one during the early timepoint and one during the late timepoint — to probe how our networks of interest were engaged at these two distinct timepoints. As compared to the early timepoint, both patient and control groups demonstrated greater motor network engagement during the late timepoint. Importantly, compared to controls, patients revealed significantly increased engagement in the motor network during both early and late timepoints, indicating motor network hyperactivity during gain anticipation.

Compared to our healthy control group, patients with fibromyalgia demonstrated significantly higher engagement in the motor network during gain anticipation. This suggests that patients with fibromyalgia have motor network hyperactivity during their experience of gain anticipation. In healthy individuals, enhanced activation within the motor cortex occurs when anticipating reward, indicating motor preparation for reward stimuli (Knutson, Fong, et al., 2001). Primary sensorimotor areas and the supplementary motor cortex also exhibit increased activation during anticipation of reward stimuli (Bjork et al., 2010; Ernst et al., 2004). Patients with chronic pain often have deficits in motor performance (Bank et al., 2015; Lamoth et al., 2006). Therefore, motor network hyperactivity as we observed in our fibromyalgia patient cohort may be potentially related to altered motor processing in fibromyalgia (e.g., restricted movement or altered motor planning).

Meanwhile, brain motor processes are critical contributors to pain processing (Chang et al., 2018), and patients with fibromyalgia demonstrate structural and functional alterations within motor cortex and supplementary motor areas (Cook et al., 2004; Gracely et al., 2002; Pacheco-Barrios et al., 2022). For example, in a study using Functional Near-Infrared Spectroscopy (fNIRS) during a fast finger tapping task, patients with fibromyalgia demonstrated abnormal task-response with significantly lower oxyhemoglobin concentration in the motor cortex compared to controls (Gentile et al., 2019). As also observed in patients with fibromyalgia, hyperexcitability of motor networks occurs during acute painful stimulation, which further indicates dysfunctional motor cortex systems in patients with fibromyalgia (Saavedra et al., 2014).

Our results are novel in that no prior neuroimaging studies have examined the brain’s motor network during performance of a reward-related (e.g., MID) task in patients with fibromyalgia. In other patient cohorts, and similar to our present results, patients with major depressive disorder demonstrate hyperactivation in brain motor regions during reward selection (Smoski et al., 2009). Given that symptoms of fibromyalgia are also typically accompanied by affective and mood disturbances (e.g., anxiety, depression), there may be a potential interaction between dysfunctional motor systems and related clinical symptoms, which in turn may relate to increased motor activity during gain anticipation in patients with fibromyalgia.

In our exploratory analyses, both patients and controls showed increased motor network engagement closer to response (i.e., late timepoints). Supporting prior findings of altered activation in the left motor cortex during gain anticipation in patients (Knutson, Fong, et al., 2001). In the present study, patients with fibromyalgia exhibited increased left motor network engagement in *both* timepoints as compared to controls. This suggests that motor network hyperactivity is affected over the whole gain anticipation phase and does not differ between early vs. late timepoints of gain anticipation.

In contrast, for the value-driven attention network, both patients and controls showed higher engagement during the early timepoint vs. the late timepoint. In other words, even within the same anticipation phase, the value-driven attention network was more engaged when participants viewed reward stimuli (i.e., “+$5” or “+$1”) compared to a fixation mark (i.e., “+”). As this suggests, during gain anticipation, attention processes may be more driven by actual viewing of reward stimuli rather than a neutral fixation mark. Monetary reward facilitates voluntary attention to task-relevant stimuli as compared to neutral stimuli (Della Libera & Chelazzi, 2009; Pessoa & Engelmann, 2010) and even to task-irrelevant stimuli, if the stimuli are previously associated with monetary reward (Mine & Saiki, 2015). For example, during a visual target search task, participants show slower reaction times when previously reward-associated stimuli are presented as a distractor, but neutral stimuli do not affect performance when presented as a distractor (Mine & Saiki, 2015). In line with such prior data, our results provide the first neural evidence of increased engagement of the value-driven attention network during gain anticipation.

In the basal-ganglia network, our results revealed higher engagement during the late timepoint (i.e., fixation presentation) in both groups. This result is in line with gradual increases in NAcc response during the anticipation phase of the MID task, which peak during fixation (i.e., after the reward stimulus “cue” has disappeared) (Martucci et al., 2018; Park, Deng, et al., 2022). Further studies are needed to verify and examine the role of the basal-ganglia networks during specific timepoints in gain anticipation.

Despite its common utility for both resting-state and task-based fMRI analysis, for the study of chronic pain, the ICA approach has been previously used mostly with resting-state functional MRI studies data. For example, in diabetic neuropathic pain patients, functional connectivity within the DMN is disrupted during rest (Cauda et al., 2009). Similarly, in patients with fibromyalgia, functional connectivity is altered within the DMN and within the right executive attention network during rest (Napadow et al., 2010). However, to our knowledge, no prior studies have investigated ICA-derived brain networks during task performance in patients with chronic pain. Meanwhile, reward and motivation are inherently important to the study of chronic pain, and require further investigation in new ways (Borsook et al., 2016; Loggia et al., 2014; Martucci et al., 2018, 2019; Porreca & Navratilova, 2017). Therefore, through this study, we sought to probe how brain network activity is altered in patients with fibromyalgia during the active engagement of brain reward processes.

In our primary analysis, we measured the strength of correlations between network timecourses for each of our three networks of interest and a gain anticipation task timecourse. The advantage of this approach is that it can reveal the level of network engagement during the task. Specifically, we used this strategy to reveal each network’s involvement in the gain anticipation aspect of the task (i.e., our focus was on how much of the network activity during task performance was driven by gain anticipation processes in the brain). Notably, the timecourse of each network reflected an individual participant’s unique representation of the network’s activity during task engagement, as these network timecourses were derived from data obtained during the individual’s participation in the task. Thus, by using a completely data-driven approach to analyze task-based fMRI data, we aimed to provide broader insights to how brain networks are altered during gain anticipation in patients with fibromyalgia.

Some limitations of our approach should be taken into account. While we limited our analysis to three reward-relevant networks of interest, none of the network timecourses were significantly correlated with the gain anticipation task timecourse. However, this result is logical and can be explained as being due to inherent differences in the data contributing to each type of timecourse -- the gain anticipation timecourse included data from *only* the gain anticipation phase of the task while the network timecourse included data from all timepoints (e.g., gain anticipation, loss anticipation, reward outcome phase). Despite this lack of significant correlation between the task and network timecourses, our study question was focused on group differences in the degree of correlation between timecourses, which indicated meaningful results and were consistent with the broader reward literature. Additionally, as medications are known to affect brain processing and networks, and can introduce variability (Martucci et al., 2019), it is a limitation that we were not able to directly measure the effect of medications taken by patients in this study. Notably, all participants in the present study were not taking opioid medications. In addition, as the majority of referred fibromyalgia patients are women, only females were recruited in this study. Because sex differences can influence on brain activity in response to rewards in patients with fibromyalgia (Baker et al., 2022; Wolfe et al., 2018), evaluating the current findings with males will provide more comprehensive insights. Despite these limitations, the present study contributes novel findings of exaggerated motor functional network responses during anticipated reward in fibromyalgia.

In conclusion, using a data-driven ICA approach to analyze reward task fMRI data, we observed significantly altered motor network engagement during gain anticipation in patients with fibromyalgia. With an exploratory analysis, our results further provide evidence of different levels of motor network engagement during early and late timepoints of the gain anticipation phase. While both patients and controls showed increased motor network engagement during the late timepoint, which supports the notion of motor preparation prior to target response, patients exhibited greater engagement of the motor network during both early and late timepoints. Building upon established brain reward-related alterations in patients with chronic pain (Baker et al., 2022; Kim et al., 2020; Loggia et al., 2014; Martucci et al., 2018, 2019; Park, Baker, et al., 2022; Park, Deng, et al., 2022), our results provide further insight to how brain networks are altered in a specific chronic pain condition, fibromyalgia, during gain anticipation. In patients with fibromyalgia, our observed motor network hyperactivity may be a function of altered behavioral motivation during gain anticipation and may relate to altered motor processing in fibromyalgia, as observed in chronic pain conditions more broadly. Our results provide new knowledge regarding how altered brain reward processes in patients with fibromyalgia relates to reward-relevant brain network function and underscore the importance of further investigation between motor and reward systems in patients with chronic pain for future therapeutic benefit.

## Data Availability

All data produced in the present study are available upon reasonable request to the authors.

## Author Contributions

The authors confirm contribution to the paper as follows: S.H.P., A.M.M., and K.T.M. were responsible for the conception and design of the study. S.H.P., A.K.B., and K.T.M. collected the data. S.H.P., A.M.M., and K.T.M. analyzed the data. S.H.P., A.M.M., A.K.B., C.L., and K.T.M. were involved in the interpretation of results. S.H.P. produced the initial manuscript draft. S.H.P., A.M.M., A.K.B., C.L., and K.T.M. edited and revised the manuscript. All authors reviewed and approved the final version of the manuscript.

## Acknowledgments

The authors express gratitude to Lindsie Boerger, Meghna Nanda, Vinit Krishna, and Sharon Norman for their valuable support in recruitment, data collection, regulatory oversight, data organization, and assistance with data quality control. We also sincerely thank the Duke University Brain Imaging and Analysis Center (BIAC), particularly, Drs. Allen Song and Todd Harshbarger, for assistance with fMRI sequence and protocol design, and Susan Music, Jennifer Graves, and Lamont Conyers for their expert assistance with MRI/fMRI data collection. Finally, we thank all of the study participants for their contribution to clinical research. The National Institutes of Health (NIH) provided funding for this study (R00 DA040154 and R01 DA055850).

