## Supplementary Figures for "Altered Functional Networks during Gain Anticipation in Fibromyalgia"

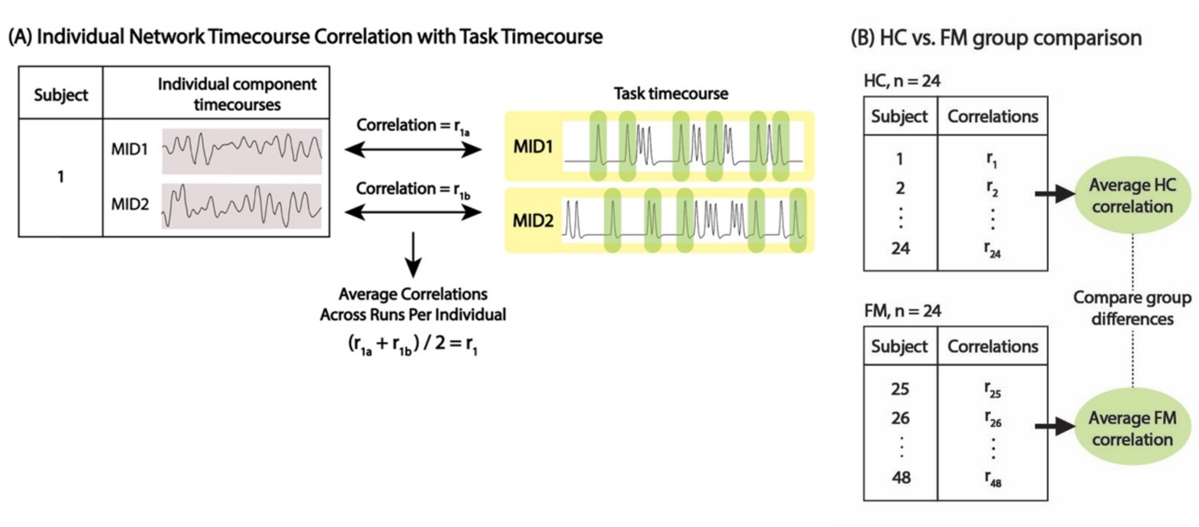


Supplementary Figure 1. Detailed Pipeline for the Analyses of Individual-Level and Group-Averaged Functional Network Correlations with the Gain Anticipation Task Timecourse.

(A) We constructed a task timecourse model (for MID1 run and MID2 run separately) by convolving the timecourse of the anticipation phase with the hemodynamic response function (using FSL). Then, we correlated the task timecourse with individual network timecourses resulting from the ICA analysis. This was conducted separately for the MID1 run and the MID2 run, for each individual participant's dataset. (B) Next, we obtained the averaged (MID1 and MID2) correlation value for each individual participant. Finally, we calculated the group-averaged correlation values to be used in the final group comparison. Abbreviation: FM, fibromyalgia; HC, healthy control; MID, monetary incentive delay; r, Pearson correlation coefficient.

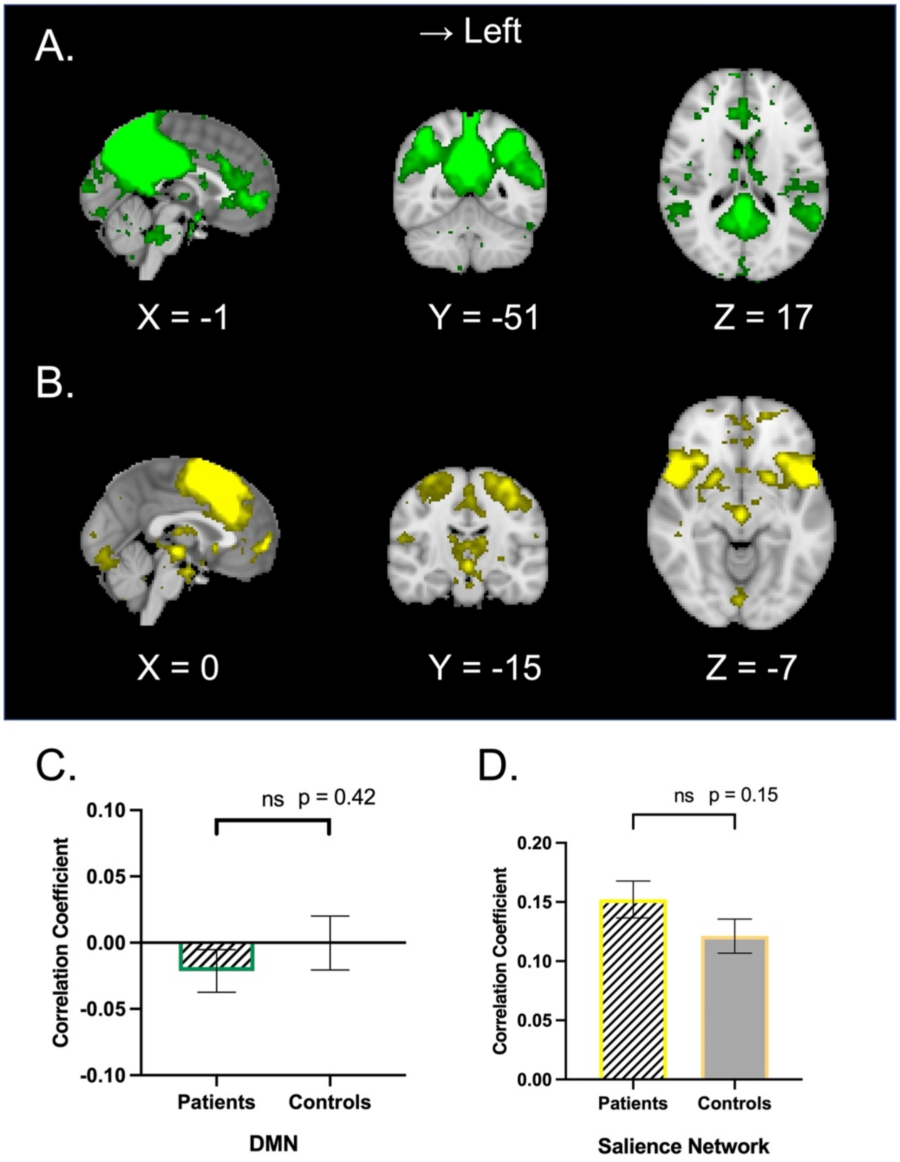


Supplementary Figure 2. Correlation Coefficients for Additional Functional Networks as Engaged by the MID Task.

Additional networks which were identified from the ICA analysis included the default mode network (DMN) and the salience network. These networks were not included in our set of three networks of interest but were analyzed in post-hoc exploratory analyses. (A) The DMN included the posterior cingulate gyrus, bilateral angular gyrus, and medial frontal gyrus. (B) The salience network included the bilateral insula, bilateral amygdala, and dorsal anterior cingulate cortex. (C) The DMN timecourse x gain anticipation task timecourse correlation coefficient was not significantly different between groups. (D) The salience network was also not significantly different between groups. Bar line colors are matched with the brain image color in Supplementary Figure 2A and 2B. Abbreviation: DMN, default mode network; ICA, independent component analysis; MID, monetary incentive delay.
